# Corpus callosum structural characteristics in very preterm children and adolescents: developmental trajectory and relationship to cognitive functioning

**DOI:** 10.1101/2021.11.02.21265740

**Authors:** Vanessa Siffredi, Maria Chiara Liverani, Dimitri Van De Ville, Lorena G. A. Freitas, Cristina Borradori Tolsa, Petra Susan Hüppi, Russia Hà-Vinh Leuchter

**Author notes:** These authors contributed equally to this work (Petra Susan Hüppi, Russia Hà-Vinh Leuchter). **Address correspondence to:** Vanessa Siffredi, Child Development Lab & Medical Image Processing Lab – Campus Biotech, Chemin des Mines 9, 1202 Genève.

## Abstract

Previous studies suggest that structural alteration of the corpus callosum, i.e., the largest white matter commissural pathway, occurs after a preterm birth in the neonatal period and lasts across development. The present study aims to unravel corpus callosum structural characteristics across childhood and adolescence in very preterm (VPT) individuals, and their associations with general intellectual, executive and socio-emotional functioning. Neuropsychological assessments, T1-weighted and multi-shell diffusion MRI were collected in 79 VPT and 46 full term controls aged 6 to 14 years. Volumetric, diffusion tensor and neurite orientation dispersion and density imaging (NODDI) measures were extracted on 7 callosal portions using TractSeg. A multivariate data-driven approach (partial least squares correlation) and an age normative modelling approach were used to explore associations between callosal characteristics and neuropsychological outcomes. The VPT and a full-term control groups showed similar trends of white-matter maturation over time, i.e., increase FA and reduced ODI, in all callosal segments, that was associated with increase in general intellectual functioning. However, using age-related normative modelling, findings show atypical pattern of callosal development in the VPT group, with reduced callosal maturation over time that was associated with poorer general intellectual and working memory functioning, as well as with lower gestational age.

**Highlights:**

- Callosal development was explored in full-term and very preterm (VPT) aged 6 to 15 years
- Neuropsychological, callosal volumetric, tensor and NODDI measures were used
- Age-related normative modelling revealed atypical callosal development in VPT
- In VPT, atypical callosal maturation was associated with poorer cognitive functioning
- In VPT, greater prematurity was associated with increased atypical callosal maturation

**CRediT roles:** Vanessa Siffredi: Conceptualization; Data curation; Formal analysis; Investigation; Methodology; Project administration; Software; Visualization; Writing - original draft; Writing - review & editing.

Maria Chiara Liverani: Data curation; Investigation; Methodology; Project administration; Writing - review & editing.

Dimitri Van De Ville: Methodology; Resources; Software; Supervision; Writing - review & editing.

Lorena G. A. Freitas: Data curation; Writing - review & editing.

Cristina Borradori Tolsa: Investigation; Project administration; Resources; Supervision; Validation; Writing - review & editing.

Petra Susan Hüppi: Conceptualization; Funding acquisition; Methodology; Project administration; Resources; Supervision; Validation; Writing - review & editing.

Russia Hà-Vinh Leuchter: Conceptualization; Investigation; Methodology; Project administration; Resources; Supervision; Validation; Writing - review & editing.

## Introduction

With more than 190 million axon fibres, the corpus callosum is a major white matter commissural pathway that connects neurons between the two cerebral hemispheres of the human brain (Edwards, Sherr, Barkovich, & Richards, 2014). The corpus callosum (CC) plays a crucial role for interhemispheric communication of low-level sensory and motor information but also for higher-level cognitive information (Gazzaniga, 2000; Hofer & Frahm, 2006; Hofer, Merboldt, Tammer, & Frahm, 2008). As a consequence, structural alteration of the corpus callosum has been associated with reduced general intellectual (Fryer et al., 2008; Hutchinson et al., 2009; E. Luders et al., 2007; Siffredi et al., 2018), executive (Brown, Panos, & Paul, 2020; Hinkley et al., 2012; Kok et al., 2014; Lyoo et al., 1996; Siffredi et al., 2017) and socio-emotional functions (Badaruddin et al., 2007; Barnea-Goraly et al., 2004; Bridgman et al., 2014; Hardan et al., 2009; Paul et al., 2007). In typical foetal development, the basic structure and shape of the corpus callosum is completed by 20 gestational weeks (Edwards et al., 2014; Lindwall, Fothergill, & Richards, 2007; Rakic & Yakovlev, 1968). However, it continues to increase in size over the third trimester of pregnancy and postnatally up until 2 years of age, when it reaches a size comparable to adults (Giedd et al., 1996; Malinger & Zakut, 1993; Pujol, Vendrell, Junque, Marti-Vilalta, & Capdevila, 1993; Rauch & Jinkins, 1994). This developmental period is accompanied by axon growth, followed by a period of synaptic pruning (Innocenti & Price, 2005). Thus, very preterm (VPT) birth, i.e., before 32 completed weeks of gestation, occurs during a highly sensitive period of callosal development.

In VPT individuals, previous studies indeed showed structural alteration of the corpus callosum in the neonatal period that lasts across childhood, adolescence and young adulthood (de Bruïne et al., 2011; Hasegawa et al., 2011; Huang, Chou, Tsao, & Chen, 2020; Hüppi et al., 1998; Teli et al., 2018; Thompson et al., 2011; van Pul et al., 2012). Firstly, reduction in callosal volume in VPT have been found at different ages across childhood from 7 years old (Rademaker et al., 2004), adolescence and young adulthood up until 20 years of age (Caldú et al., 2006; Narberhaus et al., 2007; Narberhaus et al., 2008; Nosarti et al., 2014; Nosarti et al., 2004). Volumetric alteration is found more specifically in the posterior area of the corpus callosum and reduction in callosal volume is correlated with increased prematurity (Caldú et al., 2006; Narberhaus et al., 2007; Narberhaus et al., 2008; Nosarti et al., 2004). Using longitudinal data, Allin and colleagues (2007) showed increased volumetric growth from 15 to 19-year-old in preterm individuals compared to full-term controls (Allin et al., 2007). However, despite this accelerated growth, CC volume seems to stay reduced in this population. Secondly, structural alteration of the corpus callosum has been explored using diffusion-weighted imaging (DWI), and the diffusion tensor model is known to provide insight into the microstructure and connectivity of white matter tracts (Pierpaoli, Jezzard, Basser, Barnett, & Di Chiro, 1996). A reduction in callosal fractional anisotropy (FA) values, a measure of the directionality of diffusion, was found at early ages in VPT children (de Almeida et al., 2020; Hüppi et al., 1998; Jo et al., 2012; O’Gorman et al., 2015; Thompson et al., 2015) as well as in young adolescents (Nagy et al., 2003), especially in posterior CC portions. Mean diffusivity (MD), a measure of overall diffusion (with a larger MD associated with reduced integrity of the white-matter (Thompson et al., 2011), was found to be increased in VPT during childhood (Jo et al., 2012) and young adulthood (Kontis et al., 2009). Importantly and in line with the implication of the corpus callosum in neurodevelopment, these volumetric and microstructural callosal alterations in VPT individuals have been consistently associated with poorer cognitive functioning, including general intellectual and executive outcomes (Kontis et al., 2009; Narberhaus et al., 2007; Narberhaus et al., 2008; Nosarti et al., 2004; Rademaker et al., 2004).

While the tensor model and its indices of FA and MD are commonly used to study white matter microstructural properties, they lack specificity on informing on the underlying biological mechanisms. As an illustration, a reduction of FA in a white matter tract can be driven by multiple contributing factors such as decreased myelination or decreased axonal fibre density (Jones, Knösche, & Turner, 2013). Furthermore, lower FA will be evaluated in voxels containing crossing fibres compared to those without crossing fibres, a finding which might incorrectly be interpreted as reduced structural integrity (Nemanich, Mueller, & Gillick, 2019). In this context, multi-shell diffusion imaging combined with the application of advanced statistical models provides an opportunity to measure more specific information regarding white matter microstructural properties. One such model is the neurite orientation dispersion and density imaging (NODDI; (Zhang, Schneider, Wheeler-Kingshott, & Alexander, 2012)) that captures neurite (dendrites and axons) morphology, providing parameters including neurite density index (NDI) and orientation density index (ODI). More specifically, while NDI represents the intra-cellular volume fraction, estimating the density of axons within a voxel, ODI represents the angular variation of neurite orientations, reflecting the bending and fanning of axons (Zhang et al., 2012). This model has been found to be valuable in characterising early microstructural development (Kunz et al., 2014).

Building on previous findings and using advanced methodology, this study aimed to examine corpus callosum structural development in VPT children and adolescents aged 6 to 14 years using a comprehensive set of structural and microstructural measures and its association with general intellectual, executive and socio-emotional functioning. Firstly, patterns of associations between callosal volumetric, tensor and NODDI measures, and age, gestational age and neuropsychological functioning were explored in full-term and VPT individuals. Secondly, we employed an age-related normative modelling approach on callosal volumetric, tensor and NODDI measures. This approach consists of fitting a mathematical distribution that finds the relationship between age and a given callosal structural characteristic measure, as well as the variation in this relationship expected in a group of full-term controls (Bethlehem et al., 2020; Lefebvre et al., 2018; Marquand, Rezek, Buitelaar, & Beckmann, 2016). Callosal structural characteristic measures of VPT individuals can then be understood in relation to this normative model and allows identification of deviations from normative callosal development for each individual. In the VPT group, we then used this approach to explore specific association between deviation from normative corpus callosum structural development with age, gestational age, neuropsychological functioning.

In summary, associations between corpus callosum structural development—using volumetric, tensor and NODDI measures—and age, gestational age and neuropsychological functioning, were explored in full-term and VPT individuals using both a traditional and a normative modelling approaches. This procedure allowed us to, first, establish how callosal structural development is influenced by age and gestational age factors in full-term individuals, as well as association with general intellectual, executive and socio-emotional functioning. Secondly, it allowed us to better capture atypical callosal structural development in VPT individuals and its association with general intellectual, executive and socio-emotional functioning.

## Methods

### Participants

Participants of the current study were recruited for two intervention studies (‘Mindful preterm teens’ study (Siffredi et al., 2021); and ‘Vis-à-Vis’ study), between January 2017 and July 2019. 392 VPT children and adolescents born < 32 gestational weeks between 01.01.2003 and 31.12.2012, in the Neonatal Unit at the Geneva University Hospital (Switzerland) and followed up at the Division of Child Development and Growth, were invited to participate. VPT children and adolescents were excluded if they had an intelligence quotient below 70, sensory or physical disabilities (cerebral palsy, blindness, hearing loss), or an insufficient understanding of French. A total of 108 VPT participants were enrolled. Of the 108 participants enrolled, 79 completed both the brain MRI scan and neuropsychological assessment. A total of 65 were included in the current study (diffusion sequences not completed: n=9; high level of motion artefacts: n=5). Moreover, 46 term-born children and adolescents aged between 6 and 14 years old were recruited through the community. Of the 46 participants, 41 completed both the brain MRI scan and neuropsychological assessment. A total of 39 were included in the current study (diffusion sequences not completed: n=2).

This study was approved by the Ethics Committee. Written informed consent was obtained from the principal caregiver and from the participant.

### Neuropsychological measures

Participants’ general intellectual, executive and socio-emotional functioning were assessed using neuropsychological testing and computerised neurocognitive tasks, for detailed information see Supplementary Table S1.

#### (i) General intelligence measure

In participants from 6 years to 9 years and 11 months old, the Kaufman Assessment Battery for Children – 2nd Edition (K-ABC-II; (Kaufman & Kaufman, 2013)) was used to evaluate the Fluid-Crystallized Index (FCI) as a measure of general intellectual functioning. The FCI is derived from a linear combination of 10 core subtests that composed five first-order scale scores (i.e., Short-Term memory, Long-Term Storage and Retrieval, Visual Processing, Fluid Reasoning, and Crystallized Ability). For children younger than 7 years of age, a different subset combination is administered to calculate the FCI. In participants from 10 to 14 years of age, the Wechsler Intelligence Scale for Children – 4th Edition (WISC-IV; (Wechsler, 2003) was used to evaluate the General ability index (GAI) as a measure of general intellectual functioning. The GAI is derived from the core verbal comprehension and perceptual reasoning subtests. Both of these measures of general intellectual functioning, FCI and GAI, have a mean of 100 and a standard deviation of 15.

#### (ii) Executive functioning measures

Executive functioning was assessed based on the model of Anderson (Anderson, 2002) using: a) the Letter-Number Sequencing subtest from Wechsler Intelligence Scale for Children, 4^th^ Edition (WISC-IV) assessing working memory, which belongs to the cognitive flexibility subdomain (Anderson, 2002); and b) a computerised Flanker Visual Filtering Task, in which reaction time of the congruent condition was used to assess speed of processing, which belongs to the information processing subdomain; and the inhibition score (accuracy in incongruent conditions – accuracy in congruent conditions) was used as a measure of the attentional control subdomain (Anderson, 2002; Christ, Kester, Bodner, & Miles, 2011). Given that age-related increases in executive functioning are known to exist, raw scores were regressed on age at testing; the standardised residuals for the three executive measures were retained for following analysis, called working memory, processing speed and inhibition scores.

#### (iii) Socio-emotional functioning measures

Socio-emotional functioning was assessed using subtests of the Developmental Neuropsychological Assessment - 2nd Edition (NEPSY-II; (Korkman, Kirk, & Kemp, 2007) including : a) the Affect Recognition subtest giving a total score assessing facial emotional recognition; and b) the Theory of Mind subtest giving a total score measuring the ability to understand mental contents, such as belief, intention or deception. Given age-related increases in socio-emotional functioning, raw scores were regressed on age at testing; the standardised residuals for the two socio-emotional measures were retained for following analysis, called affect recognition and theory of mind scores.

### Magnetic resonance imaging

#### Magnetic Resonance Imaging (MRI) acquisition

MRI data were acquired at the Campus Biotech in Geneva, Switzerland, using a Siemens 3T Magnetom Prisma scanner. All participants completed a simulated “mock” MRI session prior to their MRI scan. This preparation process was conducted by trained research staff and allowed participants to familiarise themselves with the scanner and the scanning process, eventually raising any concerns they might have had prior to the MRI scan. Furthermore, this process is known to facilitated acquisition of good quality MRI images in children and adolescents (de Bie et al., 2010; Tamnes, Roalf, Goddings, & Lebel, 2018). Structural T1-weighted MP-RAGE (magnetization-prepared rapid gradient-echo) sequences was acquired using the following parameters: voxel size = 0.9 × 0.9 × 0.9 mm; repetition time (TR) = 2,300 ms; echo time (TE) = 2.32 ms; inversion time (TI) = 900 ms; flip angle (FA) = 8°; and field of view (Fov) = 240 mm. A multi-shell diffusion-weighted (DW) echo planar imaging (EPI) protocol was used and included four shells. The first sequence, referred to as ‘*b200*’, was acquired with b-values of 200□s/mm2, 10 gradient directions, 4 b-value = 0□s/mm2 images, TR = 7000□ms, TE = 87□ms, FOV = 234 ×□243□mm, slice thickness = 1.3 mm, voxel size = 1.3 ×□1.3 ×□1.3 mm. The second sequence, referred to as ‘*b1700*’, was acquired with b-values of 1700□s/mm2, 30 gradient directions, 4 b-value = 0□s/mm2 images, TR = 7000□ms, TE = 87□ms, FOV = 234 ×□243□mm, slice thickness = 1.3 mm, voxel size = 1.3 ×□1.3 ×□1.3 mm. The third sequence, referred to as ‘*b4200a*’, was acquired with b-values of 4200□s/mm2, 26 gradient directions, 4 b-value = 0□s/mm2 images, TR = 7000□ms, TE = 87□ms, FOV = 234 ×□243□mm, slice thickness = 1.3 mm, voxel size = 1.3 ×□1.3 ×□1.3 mm. The forth sequence, referred to as ‘*b4200b*’, was acquired with b-values of 4200□s/mm2, 24 gradient directions, 4 b-value = 0□s/mm2 images, TR = 7000□ms, TE = 87□ms, FOV = 234 ×□243□mm, slice thickness = 1.3 mm, voxel size = 1.3 ×□1.3 ×□1.3 mm.

#### Volumetry

Volumetric measurements of the corpus callosum were based on T1-weighted MP-RAGE images and obtained using Freesurfer 5.3.0 and the recon-all function. The corpus callosum was divided into five neuroanatomically based partitions (Fischl et al., 2002). Visual quality control of the original T1 image and of the corpus callosum segmentations was completed for all participants.

#### Diffusion image preprocessing and models fitting

A flowchart summarizes diffusion image preprocessing, models fitting tractography and tractometry measures, see Figure 1.

**Figure 1.**
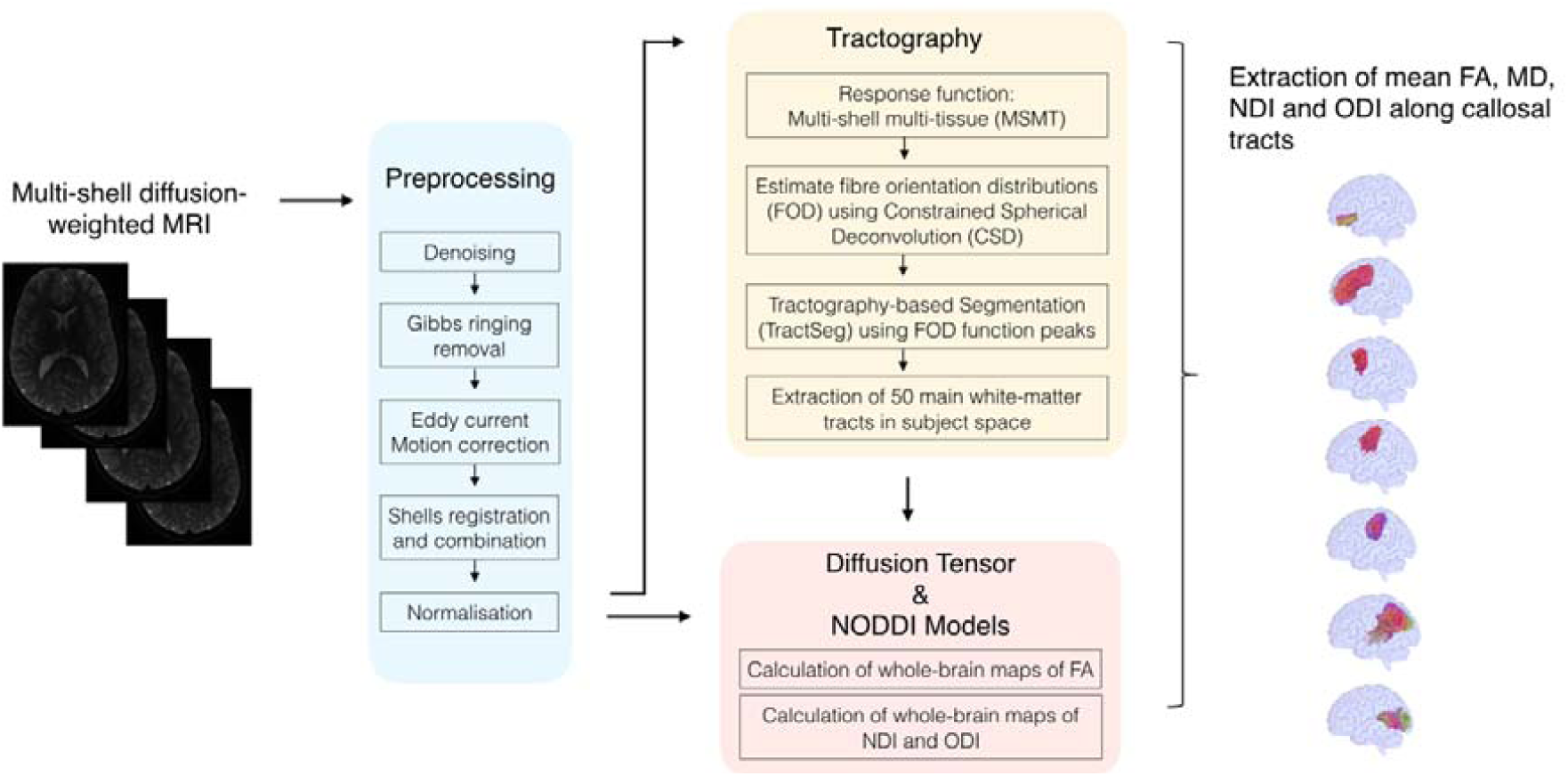
Flowchart summarizing diffusion image preprocessing, models fitting, tractography and tractometry measures. FA, Fractional Anisotropy; MD, Mean Diffusivity; NDI, Neurite Density Index; ODI, Orientation Dispersion Index

Visual inspection of raw data for brain coverage, spike artefacts, severe head motion, and other severe image artefacts was completed and participants were excluded if one of these was observed. The four diffusion shells (*b200, b1700, b4200a, b4200b*) were preprocessed independently using MRtrix3 (Tournier et al., 2019) and using the following pipeline: a) denoising (Cordero-Grande, Christiaens, Hutter, Price, & Hajnal, 2019; Veraart, Fieremans, & Novikov, 2016; Veraart, Novikov, et al., 2016), b) Gibbs ringing removal (Kellner, Dhital, Kiselev, & Reisert, 2016), c)correction for movement and eddy current-induced geometric distortions using the eddy tool implemented in FSL (Jenkinson, Beckmann, Behrens, Woolrich, & Smith, 2012). The first b =□0□s/mm2 images of the *b1700, b4200a, b4200b* sequences were linearly registered to the first b =□0□s/mm2 image of the *b200* sequence using FreeSurfer to bring them into *b200* space before merging them together. The brain extraction tool (BET) from FSL (Smith, 2002) was then applied to the combined *b200, b1700, b4200a, b4200b* image to remove non-brain tissue and subsequently intensity normalisation was applied. Following Pines and colleague’s (2020) recommendations, the resulting multi-shell diffusion weighted image was then used for models fitting, including the Diffusion Tensor Imaging (DTI) model and the Neurite Orientation Dispersion and Density Imaging (NODDI).

The DTI model was applied to the resulting multi-shell diffusion weighted image, and whole-brain maps of fractional anisotropy (FA) and mean diffusivity (MD) were calculated for each participant. FA (between 0 and 1) is a measure of the directionality of diffusion that characterise the variance of the three eigenvalues pairs that represent the direction and magnitude of diffusivity along the three orthogonal axes (*v*1, λ1; *v*2, λ2; *v*3, λ3). MD is the mean of the 3 eigenvalues and represents the average magnitude of diffusion (Tamnes et al., 2018). In addition, the NODDI Matlab Toolbox was used to extract maps of neurite density index (NDI) and fibre orientation dispersion (ODI) across the brain for each participant, http://www.nitrc.org/projects/noddi_toolbox. NDI and ODI were estimated from the resulting multi-shell diffusion weighted image using the NODDI model (Zhang et al., 2012). ODI characterises the angular variation and spatial configuration of neurite structures. NDI represents the fraction of tissue that comprises axons or dendrites (also referred to as intra-neurite volume fraction).

#### Tractography and tractometry measures

Whole-brain fibre orientation distributions (FOD) were estimated using with the multi-shell multi-tissue constrained spherical deconvolution (MSMT-CSD) method (Jeurissen, Tournier, Dhollander, Connelly, & Sijbers, 2014), resulting in a condensed representation of diffusion along three principal fibre directions per voxel according to tissue type (grey, white, cortico-spinal fluid). Tractography-based Segmentation (TractSeg) uses a supervised-learning approach with a convolutional neural network-based that directly segments tracts in the field of fibre orientation distribution function (fODF) peaks without using parcellation (Wasserthal, Neher, & Maier-Hein, 2018). TractSeg has achieved state-of-the-art performance and allows for an accurate reconstruction of fibre tracts in participant space, thus avoiding the problem of inaccurate coregistration of tracts with varying size and shape. Whole-brain fibre orientation distribution function (fODF) peaks map were input into a two stage fully convolutional neural network trained using segmented priors of 72 anatomically well-defined white matter tracts from the Human Connectome Project. The 7 segments of the corpus callosum were defined as tracts of interest. Using the tractometry function, along-tract mean FA, MD, NDI and ODI were calculated for the 7 white-matter segments of the corpus-callosum, as defined by Wasserthal and colleagues (2018) and Chandio and colleagues (2019).

### Normative Age Modelling

Age-related normative modelling was completed for all measures of corpus-callosum structural characteristics including: 5 volumetric measures, 7 along-tract corpus callosum mean FA measures, 7 along-tract corpus callosum mean MD measures, 7 mean NDI measures, 7 mean ODI measures. Age-related normative modelling was done utilising participants from the full-term control group performed, using R version 4.0.3 (R. C. Team, 2019) and RStudio version 1.3.1093 (R. Team, 2020), and adopting the methods recently described by Bethlehem and colleagues (2020). A LOESS Curve (Local Polynomial Regression) was fitted on the corpus callosum structural characteristic measures of the full-term control group. LOESS is a nonparametric method that uses local weighted regression to fit a smooth curve through points in a scatter plot. The local width of the regression (smoothing kernel) was determined by the model using the R optim function from the stats package, in which the overall smallest sum of squared errors used hyper-parameter optimisation from 5% until 100% of the full age range using Brent’s method (Brent, 1971). This approach allows to fit a potentially nonlinear relationship between age and corpus callosum structural characteristics. In the current study, age ranges from 72 to 173 months, equivalent to 6 years to 14 years and 5 months old. As a trade-off between adequate representation of developmental trajectories of corpus callosum structural characteristics and ensuring large enough subsets of full-term individuals, four age bins of 25.5 months each were created to align the full-term and the VPT groups (i.e., 72 to 96.5 months; 96.6 to 122 months; 122.1 to 148 months; 148.1 to 173 months), see Supplementary Table S2. For each age bin and every corpus callosum structural characteristic, a normative mean and standard deviation from the full-term group was calculated. These statistical norms were then used to compute a W-score (analogous to a z-score) for every VPT participant and every corpus callosum structural characteristic:

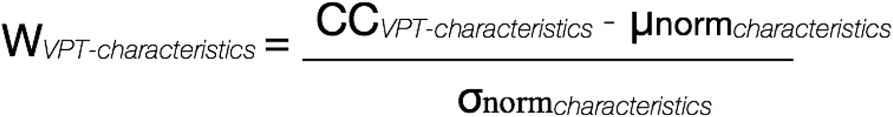

The W-score for a VPT participant quantified deviation from normative neurodevelopment for a given measure of corpus callosum structural characteristic. As W-scores are computed for every corpus callosum structural characteristics, we get a W-score for each VPT participant showing how each corpus callosum structural characteristic for that individual is atypical relative to full-term norms. See Figure 2 for a schematic overview of the age-related normative modelling procedure used here and based on Bethlehem and colleagues (2020).

**Figure 2.**
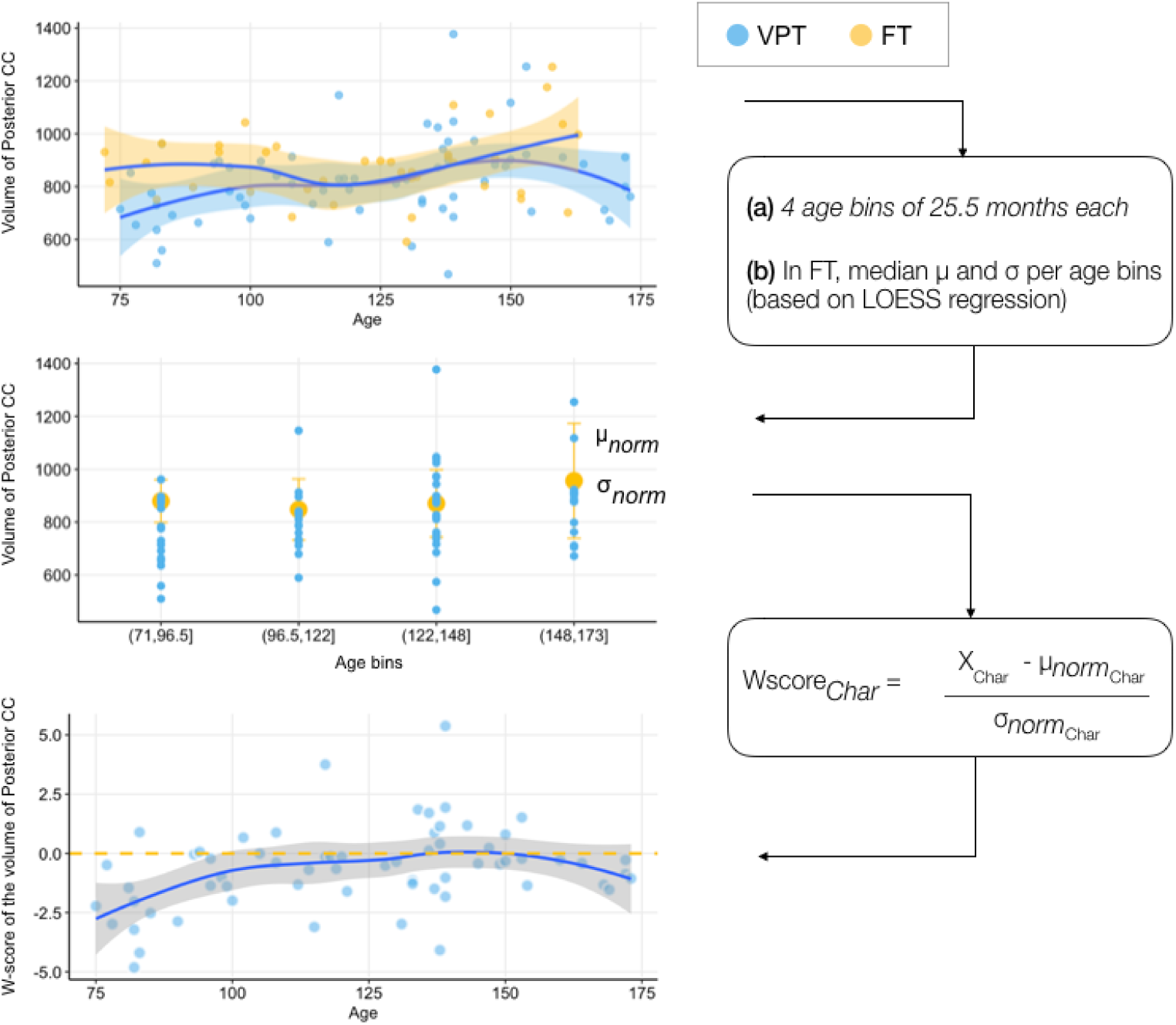
Schematic overview of the age-related normative modelling procedure based on Bethlehem and colleagues (2020). Briefly, LOESS regression was used to estimate developmental trajectories in full-term (FT) controls for each callosal measure, i.e., volumetric, tensor and NODDI measures. In Figure 2, volumetric measures of posterior portion of the corpus callosum were used. Age-bins of 25.5 months were created to align the full-term (FT) and very-preterm (VPT) groups. A normative mean and standard deviation from the full-term group were then computed for each age-bin and each callosal characteristic measure. These statistical norms were then used to compute a W-score for every VPT individuals and every callosal structural characteristic measure, called Char. VPT are represented in blue and FT in yellow.

### Statistical analyses

Partial least square correlation analyses (PLSC) were performed to evaluate association between age, gestational age, general intellectual, executive and socio-emotional functioning measures with corpus callosum structural characteristics. PLSC is a data-driven multivariate technique that maximizes the covariance between two matrices by identifying latent components which are linear combinations of the two matrices, i.e., clinical measures and corpus callosum structural characteristics measures (McIntosh & Lobaugh, 2004). A publicly available PLSC implementation in MATLAB was used: https://github.com/danizoeller/myPLS (Kebets et al., 2019; Zoller et al., 2019). Three PLSCs were computed as follow:

- In the full-term control group using corpus callosum structural characteristic measures: PLSC was used to evaluate association between age, gestational age, and general intellectual; executive; and socio-emotional functioning measures with corpus callosum structural characteristics in the full-term control group. Clinical data refers to: age, gestational age at birth and the 6 neuropsychological measures of general intellectual, executive and socio-emotional functioning. Clinical data were stored in a 39 × 8 matrix denoted X. Each row of X represents one participant and the matrix’s 8 columns consist of age, gestational age at birth and the 6 neuropsychological measures. Corpus callosum structural characteristic measures were gathered in a 39 × 33 matrix denoted Y, with each row matching one participant and each column one corpus callosum structural characteristic measure. A cross-covariance matrix was then computed between X (participants x clinical values) and Y (participants x corpus callosum structural values). Singular value decomposition was then applied to this cross-covariance matrix, resulting in latent components. Each latent component is composed of a set of clinical loadings and corpus callosum structural characteristic loadings, akin to structure coefficients. Structure coefficients lie between –1 and 1, and can be interpreted similarly to correlation values. Structure coefficients or loadings reflect the direct contribution of a specific predictor to the predictor criterion independently of others, which can be critical when predictors are highly correlated between each other (i.e., in presence of multicollinearity (Sherry & Henson, 2005)). Here, loadings indicate how strongly each clinical measures and corpus callosum structural characteristic measures contribute to the multivariate association of clinical measures and corpus callosum structural characteristics. The significance of latent components was determined by permutation testing (1000 permutations) and considered robust at p<0.01. Stability of clinical loadings and corpus callosum structural characteristic loadings were estimated using bootstrapping (500 bootstrap samples with replacement). Bootstrapped z-scores for each clinical measures and corpus callosum structural characteristic measures were calculated by dividing each clinical and corpus callosum structural characteristics correlation coefficient by its bootstrap-estimated standard deviation, and a p-value was obtained for each bootstrap’ z-score. Following the PLSC interpretation (Krishnan, Williams, McIntosh, & Abdi, 2011), the contribution of clinical loadings and corpus callosum structural characteristic loadings for a given latent component was considered robust at p < 0.01 (i.e., with a threshold of correlation coefficient above 0.4 or below -0.4).
- In the VPT group using corpus callosum structural characteristic measures: PLSC was used to evaluate association between age, gestational age, general intellectual, executive and socio-emotional functioning measures with corpus callosum structural characteristics in the VPT group. Clinical data refers to: age, gestational age at birth and the 6 neuropsychological measures of general intellectual, executive and socio-emotional functioning. The clinical data were stored in a 65 × 8 matrix denoted X. Each row of X represents one participant and the matrix’s 8 columns consist of age, gestational age at birth and the 6 neuropsychological measures. Corpus callosum structural characteristic measures were gathered in a 65 × 33 matrix denoted Y, with each row matching one participant and each column one corpus callosum structural characteristic measure. A procedure similar to the previously described PLSC conducted in full-term control was then used in the VPT group.
- In the VPT group using W-scores of corpus callosum structural characteristic: PLSC was used to evaluate association between age, gestational age, general intellectual, executive and socio-emotional functioning measures with W-scores of corpus callosum structural characteristics in the VPT group. W-scores quantify deviation from normative neurodevelopment. Clinical data refers to: age, gestational age at birth and the 6 neuropsychological measures of general intellectual, executive and socio-emotional functioning. The clinical data were stored in a 69 × 8 matrix denoted X. Each row of X represents one participant and the matrix’s 8 columns consist of age, gestational age at birth and the 6 neuropsychological measures. W-scores of corpus callosum structural characteristic measures were gathered in a 69 × 33 matrix denoted Y, with each row matching one participant and each column one W-scores of corpus callosum structural characteristic. A procedure similar to the previously described PLSC conducted on original corpus callosum measurements in VPT individuals was then used with W-scores.

For all PLSC, robust results are reported in terms of bootstrapping mean and standard deviations.

## Results

### Participant characteristics

The final sample included 65 VPT and 39 full-term participants between 6 and 14 years of age, see Table 1. Baseline characteristics were similar between VPT and full-term participants for sex and age at assessment. Socioeconomic status, as measured by the Largo score, showed group difference, with lower socio-economic status (i.e., higher Largo score) in the VPT group compared to the full-term group.

**Table 1.** Neonatal and demographic characteristics of the VPT and full-term participants

|  | VPT (n=65) | Full-term (n=39) | Group comparison |
| --- | --- | --- | --- |
| **Birth weight**<br>mean (SD) in grams | 1288.00 (378.62) | 3438.72 (412.05) | $t(95.67) = -33.89, p < 0.001$ |
| **Gestational Age**<br>mean (SD) in weeks | 29.59 (1.73) | 39.83 (1.33) | $t(74.85) = -26.56, p < 0.001$ |
| **Age at assessment**<br>mean (SD) in months | 122.97 (27.76) | 122.46 (26.55) | $t(82.98) = 0.09, p = 0.93$ |
| **Sex, n** | 33 females<br>32 males | 17 females<br>22 males | $\chi^2 (1, N = 104) = 0.5, p = 0.478$ |
| **Socio-economic risk**<br>mean (SD) | 4.44 (2.55) | 2.89 (1.29) | $t(99.01) = 4.05, p < 0.001$ |

*Note: Socio-economic status of the parents was estimated using the Largo scale, a validated 12-point score based on maternal education and paternal occupation (Largo et al., 1989). Higher largo scores reflect lower socio-economic status. Independent-sample t-test or Chi-square, as appropriate, were used to compare the VPT and the full-term groups.*

### Corpus callosum structural characteristics and association with age, gestational age and neuropsychological functioning in full-term and VPT children and adolescents

PLSC analysis applied on clinical measures (i.e., age, gestational age at birth and 6 neuropsychological measures of general intellectual, executive and socio-emotional functioning) and corpus callosum structural characteristics measures identified:

a. in the full-term control group: one statistically significant latent component, latent component 1 (p = 0.001);
b. in the VPT group: one statistically significant latent component, latent component 1 (p = 0.001).

For the two groups, a comparable latent component 1 revealed an increase in age and in general intellectual functioning associated with a general increase in mean FA and decrease in mean ODI for all segments of the corpus callosum, as well as an increase in mean NDI in the rostrum. Moreover, in the VPT group only, latent component 1 also showed a significant decrease in reaction time of the congruent condition of the flanker task (i.e., increased processing speed); and an increase in number-letter sequencing (i.e., increased working memory), associated with the same pattern of increased FA and decreased ODI for all segments of the corpus callosum, along with an increase in volume for all portions of the corpus callosum, see Figure 3. The bootstrapping mean and standard deviations are reported in Supplementary Table S3.

**Figure 3.**
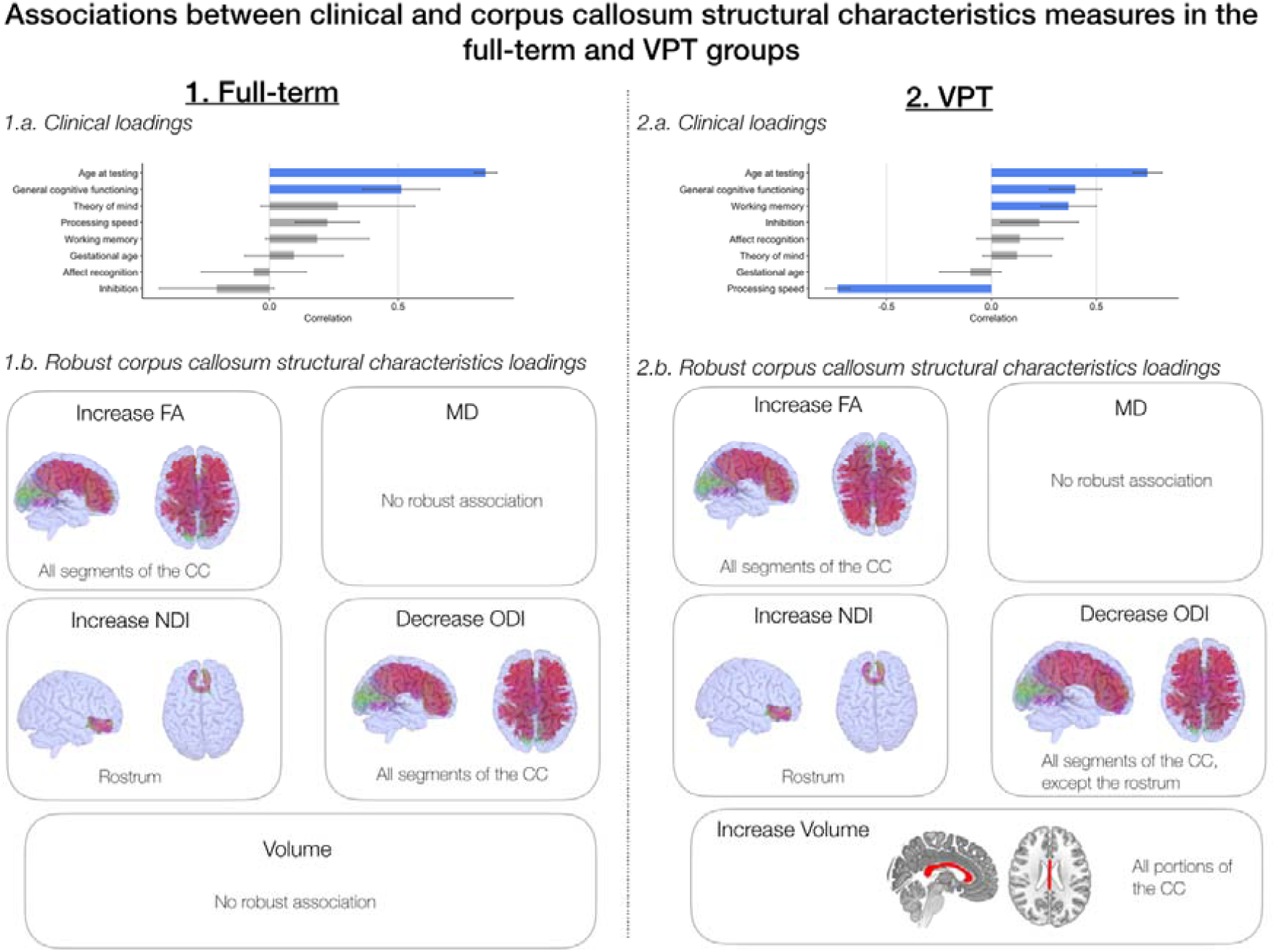
Associations between clinical and corpus callosum structural characteristics measures (i.e., volume, FA, MD, NDI, ODI) based on the PLSC analysis. 1) In the full-term control group: 1.a) Clinical loadings: the diverging graph shows mean correlations averaged across bootstrap samples and their bootstrap-estimated standard deviations (x-axis) for each clinical measure (y-axis); robust loadings are represented in blue. Of note, gestational age is in the normative range for the full-term controls. 1.b) Robust corpus callosum structural characteristics loadings for each measure and concerned corpus callosum segment or portion. 2) In the VPT group: 2.a) Clinical loadings: the diverging graph shows mean correlations averaged across bootstrap samples and their bootstrap-estimated standard deviations (x-axis) for each clinical measure (y-axis); robust loadings are represented in blue. 2.b) Robust corpus callosum structural characteristics loadings for each measure and concerned corpus callosum segment or portion.

### W-score of corpus callosum structural characteristics and association with age, gestational age and neuropsychological functioning in VPT

PLSC analysis applied on clinical measures (i.e., age, gestational age at birth and the 6 neuropsychological measures of general intellectual, executive and socio-emotional functioning) and W-scores of corpus callosum structural characteristics in the VPT group identified one statistically significant latent component: latent component 1 (p = 0.001), see Figure 4. In the VPT group, latent component 1 revealed that decreased general intellectual functioning and working memory (i.e., measured by number-letter sequencing), were associated with deviations from normative corpus callosum structural characteristics, including reduced mean FA and increased mean ODI on all segments of the corpus callosum as well as reduced MD, increased NDI and increased volume on the mid-posterior and posterior portion. Interestingly, loadings of W-scores of corpus callosum structural characteristics also show robust association with age and gestational age. Increased age at testing and decreased gestational age were overall associated with deviations from normative corpus callosum structural characteristics for mean FA and MD, (i.e., below normative expectation) and mean ODI, NDI and volumes (i.e., above normative expectation). These results reflect that the older and the “more preterm” participants are, the more they show a deviation from typical development of callosal structural characteristics. Bootstrapping mean and standard deviations are reported in Supplementary Table S4.

**Figure 4.**
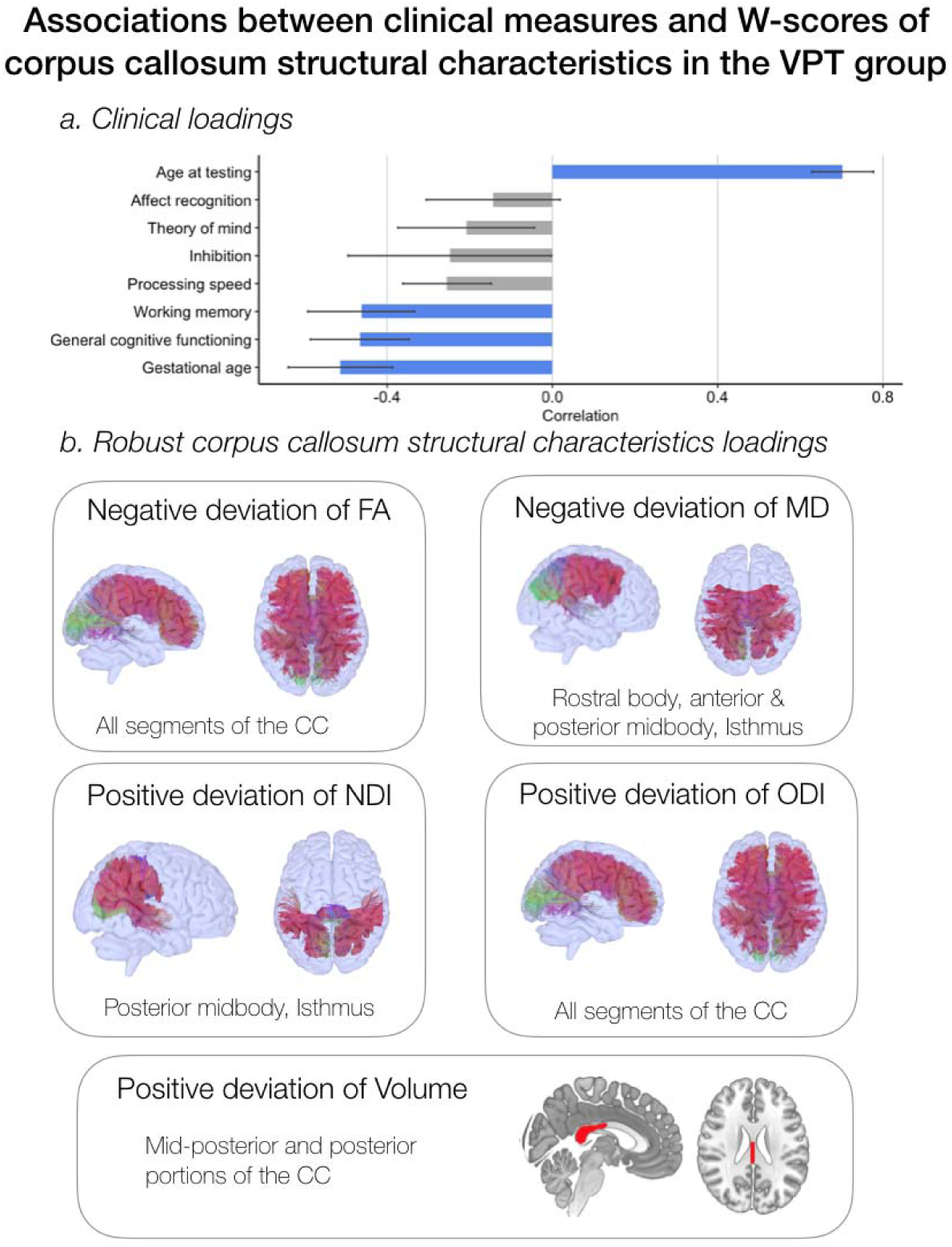
Associations between clinical measures and W-scores of corpus callosum structural characteristics measures (i.e., volume, FA, MD, NDI, ODI) based on the PLSC analysis. W-scores quantify deviation from normative neurodevelopment. a) Clinical loadings: the diverging graph shows mean correlations averaged across bootstrap samples and their bootstrap-estimated standard deviations (x-axis) for each clinical measure (y-axis); robust loadings are represented in blue. 1.b) Robust W-scores of corpus callosum structural characteristics loadings and concerned corpus callosum segment or portion.

## Discussion

The present study aims to unravel corpus callosum structural characteristics across development in VPT children and adolescents aged 6 to 14 years, as well their associations with general intellectual, executive and socio-emotional functioning.

Using multi-modal structural measures of the corpus callosum, i.e., volume, tensor and NODDI measures, the VPT and full-term control group shows an overall comparable pattern of association between age and neuropsychological functioning with corpus callosum structural characteristics. In both the VPT and full-term control groups, age and general intellectual functioning were positively associated with FA and negatively associated with ODI in most segments of the corpus callosum. During typical development, the increase in FA in callosal regions and callosal white-matter tracts is well documented within the age-range of 6 to 15 years of age (Tamnes et al., 2018; Westlye et al., 2010). For both groups, this age-related increase in FA was dominated by decreasing ODI in most callosal segment, which points to increasing coherence of axons. The rostrum also showed a specific increase in NDI, reflecting an increase in fibre diameter (Mah, Geeraert, & Lebel, 2017; Tamnes et al., 2018). This coupling of increased FA and reduced ODI in most callosal tracts was also associated with general intellectual functioning in both the VPT and the full-term control groups. These findings, reflecting the important role of the corpus callosum in general intellectual development, are in line with previous studies conducted in typically developing and VPT populations (Allin et al., 2007; Eileen Luders et al., 2007; Nosarti et al., 2004). Specific to the VPT group, age, as well as general intellectual and executive functioning (i.e., working memory and processing speed), were positively associated with the volume of all portions of the corpus callosum. Conversely, this was not found in the full-term control group. These findings suggest a late or accelerated volumetric growth of the corpus callosum in VPT school-aged children as proposed in previous studies, that reconciles with delayed maturation at earlier stages of development (Karolis et al., 2017).

Overall, in light of advanced diffusion measures of the corpus callosum, the VPT and full-term control group showed comparable trends of callosal maturation reflecting increasing axonal coherence, as well as consistent association with general intellectual functioning. Regarding volumetric measurement, only VPT children and adolescents show a specific increase in callosal volumes with age, probably reflecting a late or accelerated volumetric growth absent in typical callosal development. Moreover, in the VPT group, the pattern of increase in volume and FA and decrease in ODI was not only associated with general intellectual functioning but also with executive functioning.

Despite apparent effective growth and maturation of callosal regions and white matter tracts in the VPT group, age-related normative modelling of corpus callosum structural characteristics unravel important patterns of deviations from normative neurodevelopment. In the VPT group, age was associated with a pattern of negative deviation from normative development of FA and positive deviation of ODI on all callosal segments. This reflects that despite an apparent white-matter callosal maturation similar to the full-term with respect to increasing FA and decreasing ODI over age, callosal maturation in the VPT group is significantly slower, and the deviation from normative callosal development increases as VPT children grow older. Lower gestational age was also associated with this atypical pattern of negative deviation of FA and positive deviation of ODI; meaning that the more prematurely VPT children are born, the more they deviate from normative callosal development. These findings are consistent with previous studies showing a significant impact of the level of prematurity onto callosal structural development (Narberhaus et al., 2007; Narberhaus et al., 2008). Moreover, an atypical profile of callosal structural characteristics in VPT children was also associated with reduced general intellectual and working memory functioning. In addition to atypical developmental trajectories of FA and ODI measures for all callosal segment in VPT, posterior portions of the corpus callosum seems particularly altered in this population. Indeed, posterior portions showed positive deviation of NDI and volumes as well as negative deviation of MD associated with age. As mentioned above, these results could be interpreted as increased or accelerated maturation processes linked to increased fibre diameter and myelination (Karolis et al., 2017). Nevertheless, these neuroplastic processes that are probably in place to compensate for delayed maturation at an earlier age are not yet sufficient, as they were associated with poorer general intellectual and working memory functioning.

A limitation of the current study is the use of cross-sectional data to model callosal development. Individual variability in neurodevelopment not only occurs at inter-individual, but also intra-individual level (Foulkes & Blakemore, 2018). Therefore, characterising the factors that explain intra-individual variability during neurodevelopment is of high interest. In this context, future work should consider using longitudinal data and follow-up time points to better understand intra-individual variability.

## Conclusions

In conclusion, the full-term control and VPT groups seem to show similar trends of white-matter maturation over time, i.e., increased FA and reduced ODI in all callosal segments, that were associated with increase in general intellectual functioning in both groups. However, despite apparent growth and maturation of callosal regions and white-matter tracts in school-aged VPT similar to full-term controls, age-related normative modelling of volumetric, tensor and NODDI diffusion measures unravel atypical patterns of callosal development in the VPT population. Callosal maturation appear to deviate from normative expectation with reduced maturation over time but also with an atypical developmental trajectory. Atypical developmental trajectory of callosal maturation was associated with poorer general intellectual and working memory functioning as well as with greater prematurity. The present study illustrates how normative age modelling approach allows to shed new insight into neurodevelopmental trajectory in the VPT population and its association with functional outcomes.

## Data Availability

Ethical restrictions prevent us from making anonymised data available in a public repository. Data may be available from the Royal Children s Hospital Data Access/Ethics Committee for researchers to researchers who meet the criteria for access to confidential data by direct request to: Petra.Huppi@hcuge. There are restrictions on data related to identifying participant information and appropriate ethical approval is required prior to release. Only de-identified data will be available.

## Abbreviations

CC: Corpus Callosum
VPT: Very Preterm
DWI: Diffusion-Weighted Imaging
FA: Fractional Anisotropy
MD: Mean Diffusivity
NODDI: Neurite Orientation Dispersion and Density Imaging
NDI: Neurite Density Index
ODI: Orientation Density Index
K-ABC-II: Kaufman Assessment Battery for Children, 2nd Edition
FCI: Fluid-Crystallized Index
WISC-IV: Wechsler Intelligence Scale for Children, 4th Edition
GAI: General Ability Index
NEPSY-II: Developmental Neuropsychological Assessment, 2nd Edition
MRI: Magnetic Resonance Imaging
FOD: Fibre Orientation Distributions
MSMT-CSD: Multi-Shell Multi-Tissue Constrained Spherical Deconvolution
fODF: Fibre Orientation Distribution Function
TractSeg: Tractography-Based Segmentation
LOESS Curve: Local Polynomial Regression Curve
PLSC: Partial Least Square Correlation

## Acknowledgements

We thank and acknowledge all participating young adolescents and families who made this research possible. We also thank the Fondation Campus Biotech Geneva (FCBG), a foundation of the Swiss Federal Institute of Technology Lausanne (EPFL), the University of Geneva (UniGe), and the University Hospitals of Geneva (HUG); the Research Platform of the University Hospitals of Geneva (HUG) for their practical help.

