## Supplementary Materials for "Corpus callosum structural characteristics in very preterm children and adolescents: developmental trajectory and relationship to cognitive functioning"

### Supplementary Material

**Supplementary Table S1**. Details of the neuropsychological measures and scores

| Domains | Modalities | Measures | Description | Scores |
|---|---|---|---|---|
| General cognitive functioning | | | | |
| | Neuropsychological tests | | | |
| | | Kaufman Assessment Battery for Children – 2nd Edition (K-ABC-II; Kaufman & Kaufman, 2013) & Wechsler Intelligence Scale for Children – 4th Edition (WISC-IV; Wechsler, 2003) | | |
| | | | In participants from 6-year-old to 9 years and 11-month-old, the Kaufman Assessment Battery for Children – 2nd Edition (K-ABC-II; (Kaufman & Kaufman, 2013)) was used to evaluate the Fluid-Crystallized Index (FCI) as a measure of general intellectual functioning. The FCI is derived from a linear combination of 10 core subtests that composed five first-order scale scores (i.e., Short-Term memory, Long-Term Storage and Retrieval, Visual Processing, Fluid Reasoning, and Crystallized Ability). For children younger than 7 year of age, different subset combination is administer to calculate the FCI. In participants from 10 to 14 years of age, the Wechsler Intelligence Scale for Children – 4th Edition (WISC-IV; (Wechsler, 2003) was used to evaluate the General ability index (GAI) as a measure of general intellectual functioning. The GAI is derived from the core verbal comprehension and perceptual reasoning subtests. Both of these measures of general intellectual functioning, FCI and GAI, have a mean of 100 and a standard deviation of 15. Higher scores reflect higher general cognitive functioning. | General cognitive functioning |
| Executive competences | | | | |
| | Neuropsychological tests | | | |
| | | Letter-Number Sequencing (WISC-IV, Wechsler (2003)) | | |
| | | | The letter-number sequencing is a working memory task. Sequences of number and letters are read to the participant, and he/she is then asked to re-sequence the numbers in numerical order from lowest to highest and then to sequence the letters in alphabetical order. Raw scores were regressed on age at testing; the standardised residuals were used as a scores of working memory. Higher standardised residuals reflect higher working memory skills. | Working memory |
| | Neurocognitive computerised tasks | | | |

| | | | |
|---|---|---|---|
| | | Flanker Visual Filtering Task (Christ, Kester, Bodner, & Miles, 2011) | |
| | | The Flanker Visual Filtering Task was used to assess attentional control and information processing speed. Each trial showed a horizontal row of five fish. The participant was asked to respond as quickly as possible to whether the central fish was facing to the left or right. Congruent trials were the ones with all five fish in the horizontal row pointing in the same direction and incongruent trials were the ones with the four distracting fishes pointing in the opposite direction of the central target fish. Reaction time of the congruent condition was used to assess speed of processing, which belongs to the information processing subdomain; and the inhibition score (accuracy in incongruent conditions – accuracy in congruent conditions) was used as a measure of the attentional control subdomain. Raw scores were regressed on age at testing; the standardised residuals were used as a scores of processing speed and scores of inhibition. Higher standardised residuals reflect slower processing speed and increased difficulties in attentional control. | - Processing speed<br>-Inhibition |
| Socio-emotional competences | | | |
| | Neuropsychological tests | | |
| | | Affect Recognition (NEPSY-II, Korkman, Kirk, and Kemp (2007) | |
| | | The affect recognition subtest assesses the ability to recognise facial emotional expressions (happy, sad, anger, fear, disgust, and neutral) from photographs of children's faces in several matching tasks. In the first task, the participant selected one of the four faces that depicted the same emotion as a child's face at the top of the page. In a second task, the participant selected two photographs of faces that displayed the same affect from a selection of four photographs. Finally, the participant examined a photograph of a child's face for 5 seconds, and then from memory, selected two photographs that matched the same emotion as the face previously shown. Raw scores were regressed on age at testing; the standardised residuals were used as a scores of affect recognition. Higher standardised residuals reflect better affect recognition skills. | Affect recognition |
| | | Theory of Mind (NEPSY-II, Korkman et al. (2007)) | |
| | | The theory of mind subtest measures understanding of mental contents and other people's perspectives.<br>In the first task, questions are asked to the participant about different verbal scenarios measuring understanding of beliefs, intentions, others' thoughts, ideas and comprehension of figurative language. In the second task, participants have to match facial emotional expressions, from photographs of children's faces, to a scenario. The total raw scores were regressed on age at testing; the standardised residuals were | Theory of mind |

| | | |
|---|---|---|
| | | used as a scores of theory of mind. Higher standardised residuals reflect better theory of mind capacities. |

**Supplementary Table S2.** Description of the 4 age bins of 25.5 months created to align the full-term and the VPT groups

| Group | Age bins (in months) | Number of participants |
|---|---|---|
| VPT | (71,96.5] | 15 |
| VPT | (96.5,122] | 17 |
| VPT | (122,148] | 20 |
| VPT | (148,173] | 13 |
| FT | (71,96.5] | 8 |
| FT | (96.5,122] | 9 |
| FT | (122,148] | 15 |
| FT | (148,173] | 7 |

**Supplementary Table S3.** Bootstrapping mean and standard deviations of the loadings for neurobehavioural and callosal structural characteristics measures of the PLSC analyses in the full-term control and VPT groups.

| **Full-term control group** | | | |
|---|---|---|---|
| | **Neurobehavioural loadings** | | |
| | | **Measures** | **Loadings, bootstrapping mean (bootstrapping standard deviation)** |
| | | Gestational age | 0.095 (0.191) |
| | | Age at testing* | 0.84 (0.044) |
| | | General cognitive functioning* | 0.512 (0.149) |
| | | Inhibition | -0.205 (0.223) |
| | | Processing speed | 0.225 (0.124) |
| | | Theory of mind | 0.266 (0.299) |
| | | Affect recognition | -0.061 (0.204) |
| | | Working memory | 0.185 (0.201) |

| | | | |
|---|---|---|---|
| | **Corpus callosum structural characteristics loadings** | | |
| | | **Measures** | **Loadings, bootstrapping mean (bootstrapping standard deviation)** |
| | | Volume CC Anterior | -0.036 (0.114) |
| | | Volume CC Mid-Anterior | 0.197 (0.159) |
| | | Volume CC Central | 0.24 (0.142) |
| | | Volume CC Mid-Posterior | 0.207 (0.135) |
| | | Volume CC Posterior | 0.083 (0.12) |
| | | FA CC1* | 0.854 (0.056) |
| | | FA CC2* | 0.925 (0.03) |
| | | FA CC3* | 0.922 (0.04) |
| | | FA CC4* | 0.94 (0.026) |
| | | FA CC5* | 0.922 (0.024) |
| | | FA CC6* | 0.931 (0.022) |
| | | FA CC7* | 0.807 (0.065) |
| | | MD CC1 | 0.128 (0.324) |
| | | MD CC2 | -0.066 (0.343) |
| | | MD CC3 | 0.263 (0.325) |
| | | MD CC4 | 0.235 (0.383) |
| | | MD CC5 | 0.257 (0.332) |
| | | MD CC6 | 0.259 (0.32) |
| | | MD CC7 | 0.118 (0.229) |
| | | NDI CC1* | 0.719 (0.118) |
| | | NDI CC2 | 0.33 (0.194) |
| | | NDI CC3 | 0.155 (0.253) |
| | | NDI CC4 | 0.089 (0.245) |
| | | NDI CC5 | 0.05 (0.253) |
| | | NDI CC6 | 0.084 (0.261) |
| | | NDI CC7 | 0.271 (0.184) |
| | | ODI CC1* | -0.597 (0.151) |
| | | ODI CC2* | -0.739 (0.1) |
| | | ODI CC3* | -0.762 (0.105) |
| | | ODI CC4* | -0.821 (0.087) |
| | | ODI CC5* | -0.826 (0.07) |
| | | ODI CC6* | -0.852 (0.046) |
| | | ODI CC7* | -0.706 (0.079) |

| VPT group | | | |
|---|---|---|---|
| | Neurobehavioural loadings | | |
| | | **Measures** | **Loadings, bootstrapping mean (bootstrapping standard deviation)** |
| | | Gestational age | -0.1 (0.146) |
| | | Age at testing* | 0.743 (0.068) |
| | | General cognitive functioning* | 0.4 (0.125) |
| | | Inhibition | 0.229 (0.185) |
| | | Processing speed* | -0.733 (0.058) |
| | | Theory of mind | 0.122 (0.163) |
| | | Affect recognition | 0.136 (0.205) |
| | | Working memory* | 0.368 (0.132) |
| | Corpus callosum structural characteristics loadings | | |
| | | **Measures** | **Loadings, bootstrapping mean (bootstrapping standard deviation)** |
| | | Volume CC Anterior* | 0.24 (0.102) |
| | | Volume CC Mid-Anterior* | 0.471 (0.069) |
| | | Volume CC Central* | 0.364 (0.073) |
| | | Volume CC Mid-Posterior* | 0.179 (0.095) |
| | | Volume CC Posterior* | 0.293 (0.095) |
| | | FA CC1* | 0.605 (0.145) |
| | | FA CC2* | 0.863 (0.031) |
| | | FA CC3* | 0.805 (0.073) |
| | | FA CC4* | 0.703 (0.186) |
| | | FA CC5* | 0.904 (0.024) |
| | | FA CC6* | 0.863 (0.029) |
| | | FA CC7* | 0.715 (0.081) |
| | | MD CC1 | 0.01 (0.127) |
| | | MD CC2 | 0.421 (0.277) |
| | | MD CC3 | 0.468 (0.23) |
| | | MD CC4 | 0.456 (0.237) |
| | | MD CC5 | 0.455 (0.242) |
| | | MD CC6 | 0.375 (0.326) |
| | | MD CC7 | 0.001 (0.185) |
| | | NDI CC1* | 0.276 (0.107) |
| | | NDI CC2 | -0.123 (0.169) |
| | | NDI CC3 | 0.015 (0.167) |

| | | | |
|---|---|---|---|
| | | NDI CC4 | -0.268 (0.13) |
| | | NDI CC5 | -0.312 (0.137) |
| | | NDI CC6 | -0.275 (0.156) |
| | | NDI CC7 | 0.211 (0.134) |
| | | ODI CC1 | -0.354 (0.237) |
| | | ODI CC2* | -0.785 (0.051) |
| | | ODI CC3* | -0.561 (0.229) |
| | | ODI CC4* | -0.687 (0.131) |
| | | ODI CC5* | -0.814 (0.056) |
| | | ODI CC6* | -0.76 (0.081) |
| | | ODI CC7* | -0.673 (0.111) |

*Note: CC white-matter portion from 1 to 7 correspond to portions as defined by the Tractography-based Segmentation (TractSeg) software (Wasserthal, Neher, & Maier-Hein, 2018): Rostrum (CC1), Genu (CC2), Rostral body (CC3), Anterior midbody (CC4), Posterior midbody (CC5), Isthmus (CC6), Splenium (CC7)). * indicates significant loadings*

**Supplementary Table S4.** Bootstrapping mean and standard deviations of the loadings for neurobehavioural and W-scores of callosal structural characteristics measures of the PLSC analyses in the VPT group.

| **VPT group, using W-scores** | | | |
|---|---|---|---|
| | **Neurobehavioural loadings** | | |
| | | **Measures** | **Loadings, bootstrapping mean (bootstrapping standard deviation)** |
| | | Gestational age* | -0.514 (0.126) |
| | | Age at testing* | 0.702 (0.075) |
| | | General cognitive functioning* | -0.466 (0.12) |
| | | Inhibition | -0.248 (0.246) |
| | | Processing speed | -0.255 (0.107) |
| | | Theory of mind | -0.209 (0.165) |
| | | Affect recognition | -0.143 (0.162) |
| | | Working memory* | -0.463 (0.129) |
| | **Corpus callosum structural characteristics loadings** | | |
| | | **Measures** | **Loadings, bootstrapping mean (bootstrapping standard deviation)** |
| | | Volume CC Anterior | 0.087 (0.107) |

| | | | |
|---|---|---|---|
| | | Volume CC Mid-Anterior | 0.069 (0.136) |
| | | Volume CC Central | 0.169 (0.117) |
| | | Volume CC Mid-Posterior* | 0.206 (0.099) |
| | | Volume CC Posterior* | 0.173 (0.093) |
| | | FA CC1* | -0.562 (0.121) |
| | | FA CC2* | -0.836 (0.045) |
| | | FA CC3* | -0.702 (0.113) |
| | | FA CC4* | -0.751 (0.146) |
| | | FA CC5* | -0.916 (0.029) |
| | | FA CC6* | -0.878 (0.041) |
| | | FA CC7* | -0.692 (0.087) |
| | | MD CC1 | -0.091 (0.104) |
| | | MD CC2 | -0.071 (0.222) |
| | | MD CC3* | -0.372 (0.09) |
| | | MD CC4* | -0.311 (0.155) |
| | | MD CC5* | -0.463 (0.102) |
| | | MD CC6* | -0.418 (0.125) |
| | | MD CC7 | 0.029 (0.231) |
| | | NDI CC1 | -0.244 (0.19) |
| | | NDI CC2 | 0.2 (0.147) |
| | | NDI CC3 | 0.25 (0.12) |
| | | NDI CC4 | 0.226 (0.156) |
| | | NDI CC5* | 0.472 (0.121) |
| | | NDI CC6* | 0.389 (0.135) |
| | | NDI CC7 | 0.084 (0.161) |
| | | ODI CC1* | 0.555 (0.082) |
| | | ODI CC2* | 0.747 (0.055) |
| | | ODI CC3* | 0.635 (0.083) |
| | | ODI CC4* | 0.656 (0.158) |
| | | ODI CC5* | 0.433 (0.082) |
| | | ODI CC6* | 0.895 (0.036) |
| | | ODI CC7* | 0.596 (0.109) |

*Note: CC white-matter portion from 1 to 7 correspond to portions as defined by the Tractography-based Segmentation (TractSeg) software (Wasserthal, Neher, & Maier-Hein, 2018): Rostrum (CC1), Genu (CC2), Rostral body (CC3), Anterior midbody (CC4), Posterior midbody (CC5), Isthmus (CC6), Splenium (CC7)). * indicates significant loadings*
